# Local Mean and Pattern Standard Deviation Map for Disease Staging in Glaucoma

**DOI:** 10.1101/2023.09.13.23295369

**Authors:** Dennis C. Mock

## Abstract

For monitoring disease progression in glaucoma, perimetric measurements as global indices such as the mean deviation and standard pattern deviation for the visual field perimetry do not maintain a consistent diagnostic sensitivity over the entire data range. Here an analytical approach that assumes an underlying Gaussian mixture model describing the normal visual field offers an alternative solution to this situation.

---

Glaucoma is a multi-factorial eye disease that involves the degeneration of the optic nerve which leads to blindness. For this disorder, the visual function deteriorates mainly due to the death of the retinal ganglion cells and of the axons surrounding the optic nerve. It is known that particular measurements such as the mean deviation (MD) and standard pattern deviation (PSD) for the visual field (VF) perimetry do not maintain a consistent monotonic diagnostic sensitivity over the entire VF range (e.g. -33 to 0 [dB]) when monitoring the disease progression in glaucoma (1)(2)(3). Specifically, the MD is less sensitive for detecting disease related changes in the VF observed at the earlier stages (e.g. pre-perimetric, -2 to 0 [dB]), with the PSD similarly becoming less sensitive as an indicator for VF changes seen in the later stages of the disease progression (e.g. MD less than -15 dB).

There are many technical and statistical issues why this happens, leading to the standard paradigms for how the MD and PSD are used as statistical classifiers for glaucoma (4). Data analyses to reformulate the VF location sensitivity values as readout parameters like the pattern deviation (PD) and general height (GH) into separate VF spatial components may directly address some of these issues (5). For these reasons, the MD and PSD are considered as VF summary indices not intended generally for diagnosis but simply for disease staging (6).

As displayed in Figure 1, the scatterplot from the University of Washington VF supplemental dataset on GITHUB shows this common relationship for the PSD vs MD (MTD) from a large dataset (7, 8). Here, as the visual field damage progresses, as indicated with the MD decreasing, a reduction in diagnostic sensitivity is observed with the PSD value eventually declining after initially increasing with disease progression (3).

**Fig. 1.**
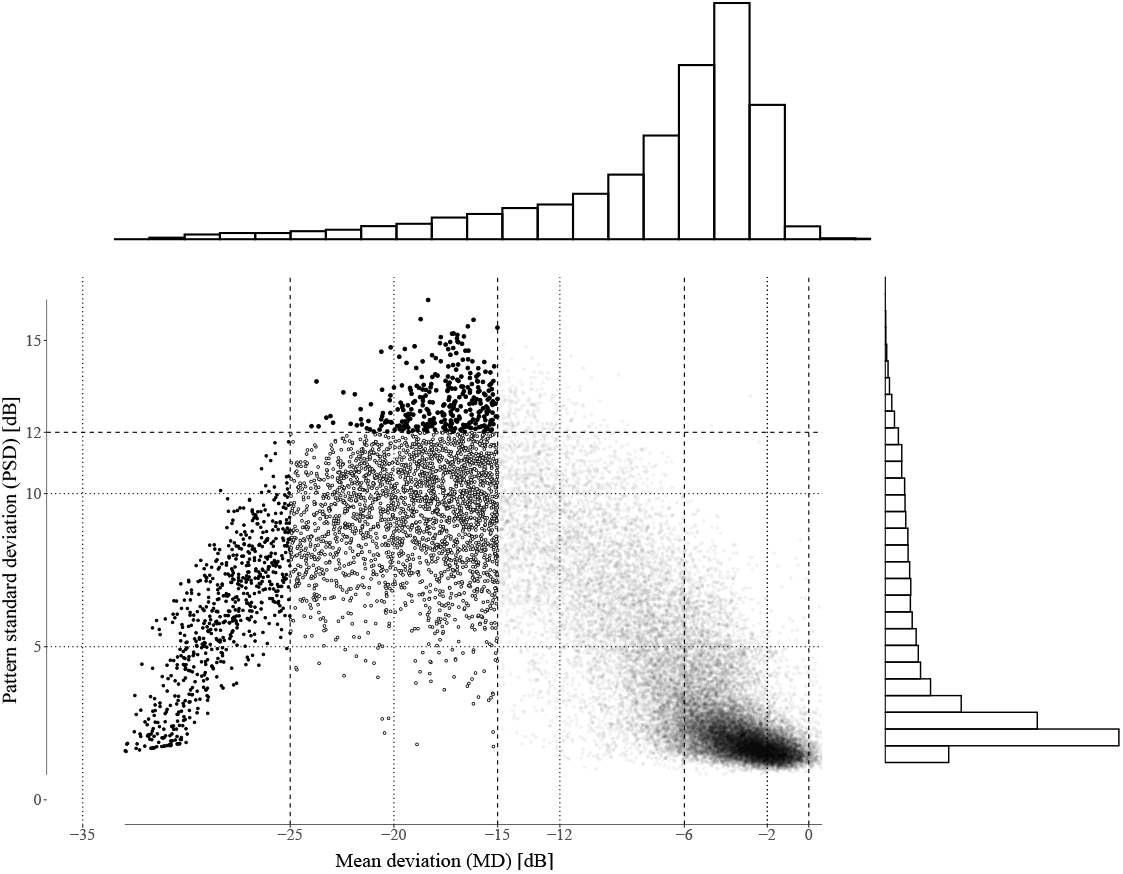
The plot displaying the pattern standard (PSD) vs mean (MD) deviation (n=28,943) from the U.W. glaucoma perimetry dataset. The VF subsets are indicated by the symbols: `-25<MD≤-15,12≤PSD[dB] ○ -25<MD≤-15,12>PSD[dB] • MD≤-25[dB] MD>-15[dB]`

Given the evidence that the functional information derived from VF perimetry corresponds to the structural abnormalities between matching *spatial* regions, recomposing the global indices MD, PSD as local, spatial indices may provide more relevant readouts for staging the disease for glaucoma (9, 10).

For instance, suppose one defines the spatial regions of an individual VF with the Garway-Heath (GH) sectors (11, 12). Then by calculating the mean (*MD*_sec_) and pattern standard deviation (*P SD*_sec_) for each of the GH sectors, the VF summary statistics (*MD*_loc_, *P SD*_loc_) for each VF is composed of separate components with its corresponding individual *MD*_sec_ and *P SD*_sec_ for each VF. Subsequently the components may be combined, for example, or summed as a total summary statistic, *MD*_loc_ and *P SD*_loc_, respectively.

For example, new summary indices are, for the *local* mean deviation:

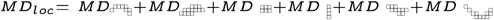

and for the *local* pattern standard deviation:

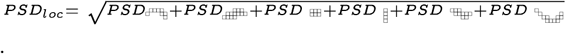

Given the original statistical formula for the MD for a single VF is defined as (13):

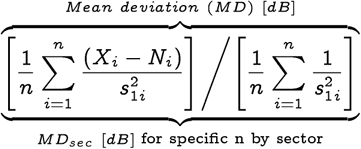

the statistical formula for calculating the new *MD*_loc_ is now

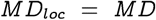

since the calculation is mathematically linear with the “mean of the sums” being equivalent to the “sum of the means” for each individual VF.

However, the statistical derivation of the *P SD*_loc_ shows the mathematical equation is non-linear and depends on the values of the local *MD*_sec_ (13). Therefore the sum of the TD components relative to a new calculation of the *MD*_sec_ would yield a different final summation value for *P SD*_loc_.

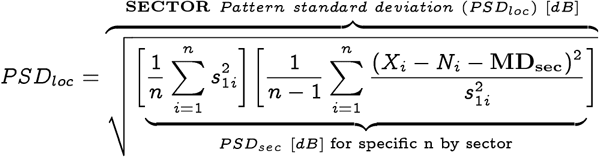

Therefore

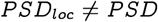

and plotting the *P SD*_loc_ value vs the MD (or *MD*_loc_) does not diminish as the MD becomes increasingly more negative (see Figure 2). Statistically, this may be understood by examining the distributions of the location sensitivities from a normal perimetry database on the individual GH sectors (see Figure 3). The overlapping Gaussian normal sub-distribution for each GH sector have varying statistical parameters, N(*μ, σ*-^2^). Therefore a VF statistical model may be represented here as a Gaussian mixture model with the varying *MD*_sec_ for each GH sector (14–16). This would computationally account for an increase in detecting local changes for the individual *P SD*_sec_ as one would expect from calculating the VF location sensitivities from the local sector rather than global VF means.

**Fig. 2.**
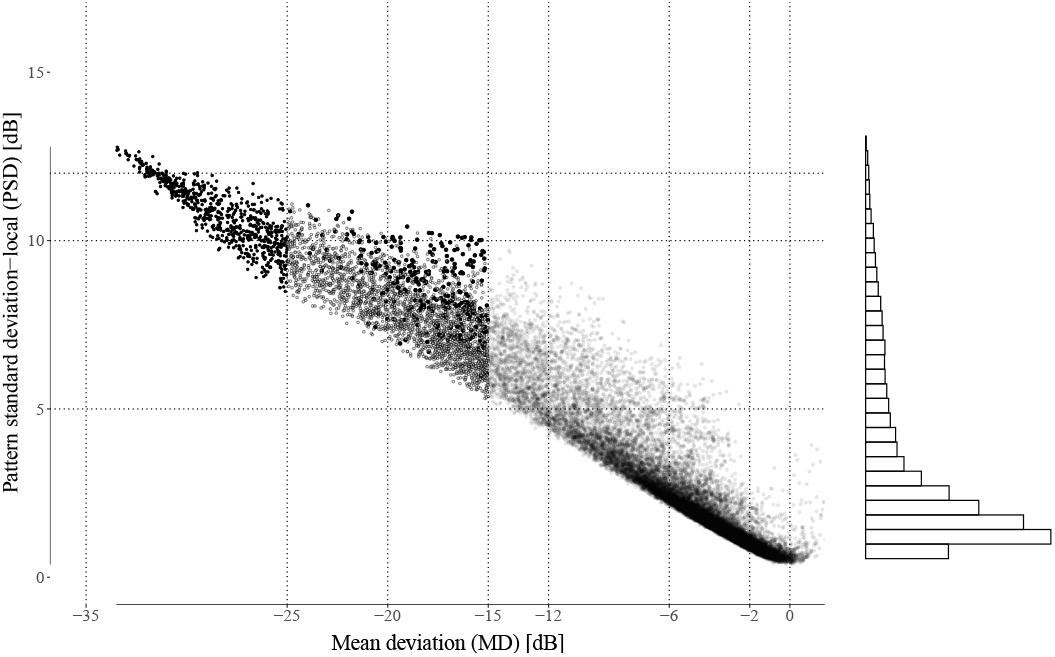
The plot displaying the pattern standard (*P SD*_loc_) vs mean(M D) deviation (n=28,943) from the U.W. glaucoma perimetry dataset. The VF subsets are indicated by the symbols: `-25<MD≤-15,12≤PSD[dB] MD>-15[dB] ∘-25<MD≤-15,12>PSD[dB] • MD≤-25[dB] MD>-15[dB]`

**Fig. 3.**
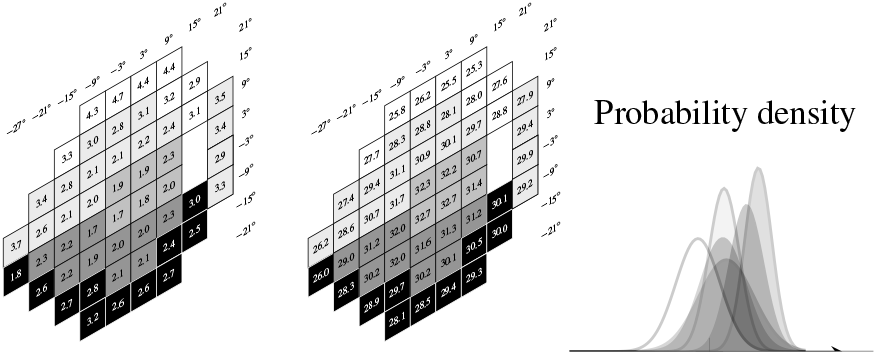
The standard deviation and normal mean thresholds, resp. with values from the literature (17) (left,middle). A graph of a Gaussian mixture model based on the normal means and standard deviations by GH sector. (right)

The revised statistical formulas for the *MD*_sec_ and *P SD*_sec_ for each Garway-Heath sector are shown pictorially in Table 1 with the individual standard deviations for the VF locations obtained from the literature (17).

**Table 1.**
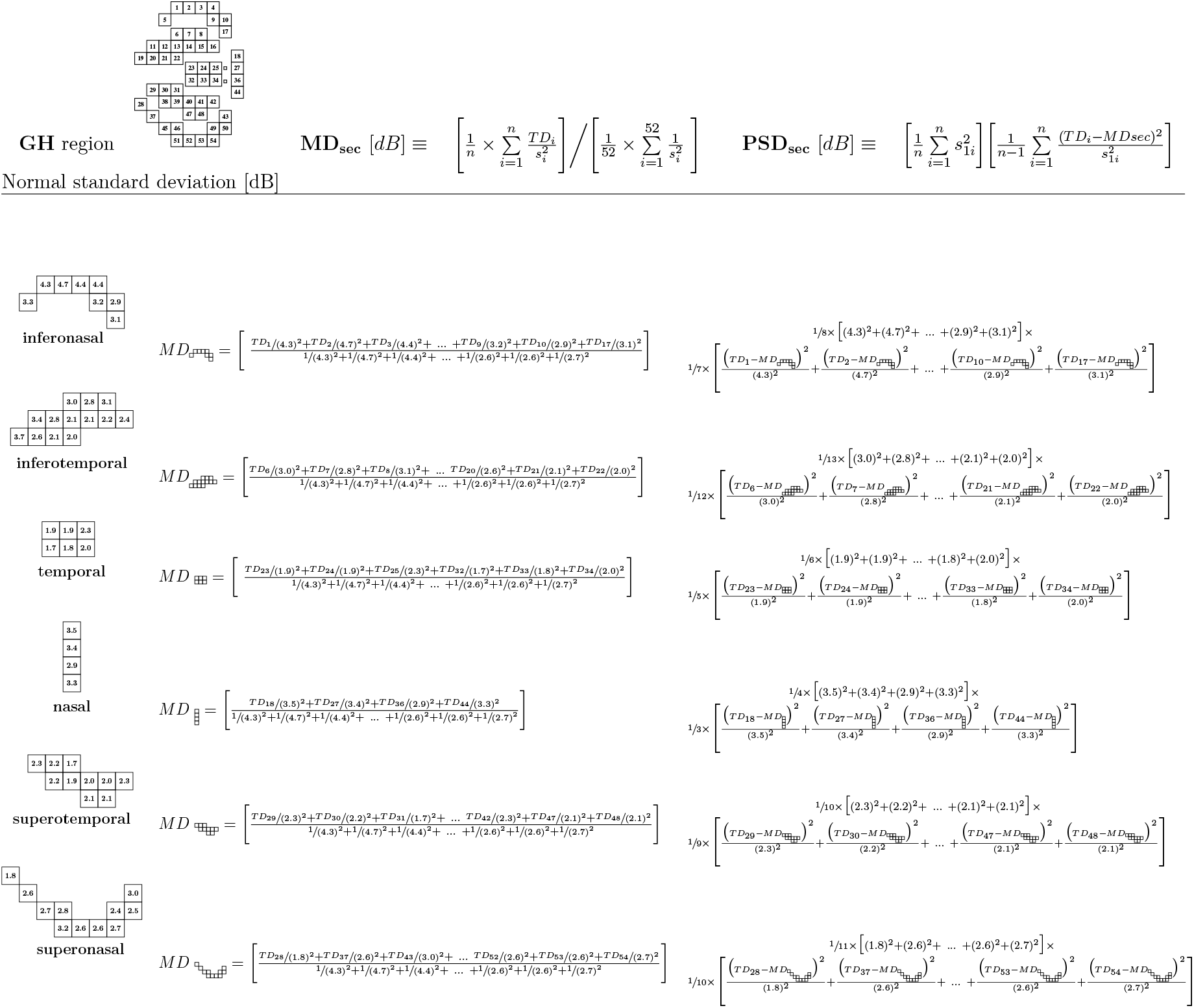
The sectors (Garway-Heath) with the standard deviation for normal variability each VF location (24-2) (left) with the corresponding calculations for the *MD*_Sec_ and *P SD*_Sec_ (middle, right), resp.(17).

As seen in Figure 2, it appears the *P SD*_loc_ retains the progression sensitivity throughout the entire perimetric range (18). This is most apparent comparing the two plots at the early (0 > MD >-6[dB]) and advanced (MD <-25[dB]) range. The information gained from remapping the *MD*_loc_ with the sum of the local pattern standard deviations *P SD*_loc_ from sectors (*P SD*_sec_) needs further investigation as now the criteria for staging disease subgroups from the *P SD*_loc_ has changed. Finally though visual inspection suggests these regional derived summary indices may offer additional information for the structure function of the progression for glaucoma (12), the traditional global summary statistics still maintain their importance for the overall baseline functional assessment (19, 20).

## Data Availability

All data produced in the present study are available upon reasonable request to the authors

https://github.com/uw-biomedical-ml/uwhvf

