## Supplemental file for "Local Mean and Pattern Standard Deviation Map for Disease Staging in Glaucoma"

**Dennis C. Mock**[a]

[a]David Geffen UCLA School of Medicine, University of California, Los Angeles, 10833 Le Conte Ave, Los Angeles, CA 90095

DRAFT

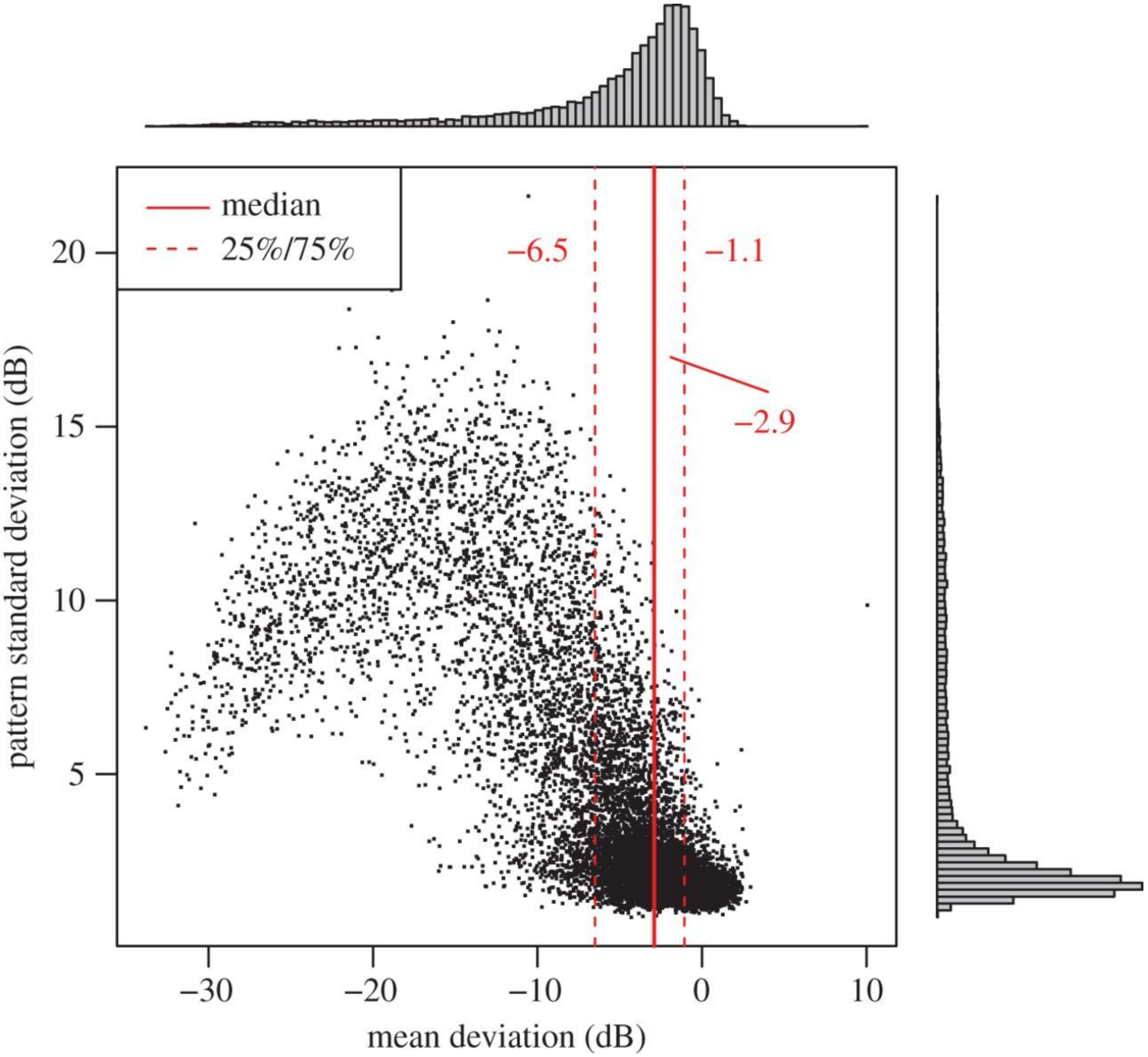


**Fig. 1.** This is a copy from Elze et.al. Figure 4. It shows the mean deviations versus pattern standard deviations of all VF measurements (1)

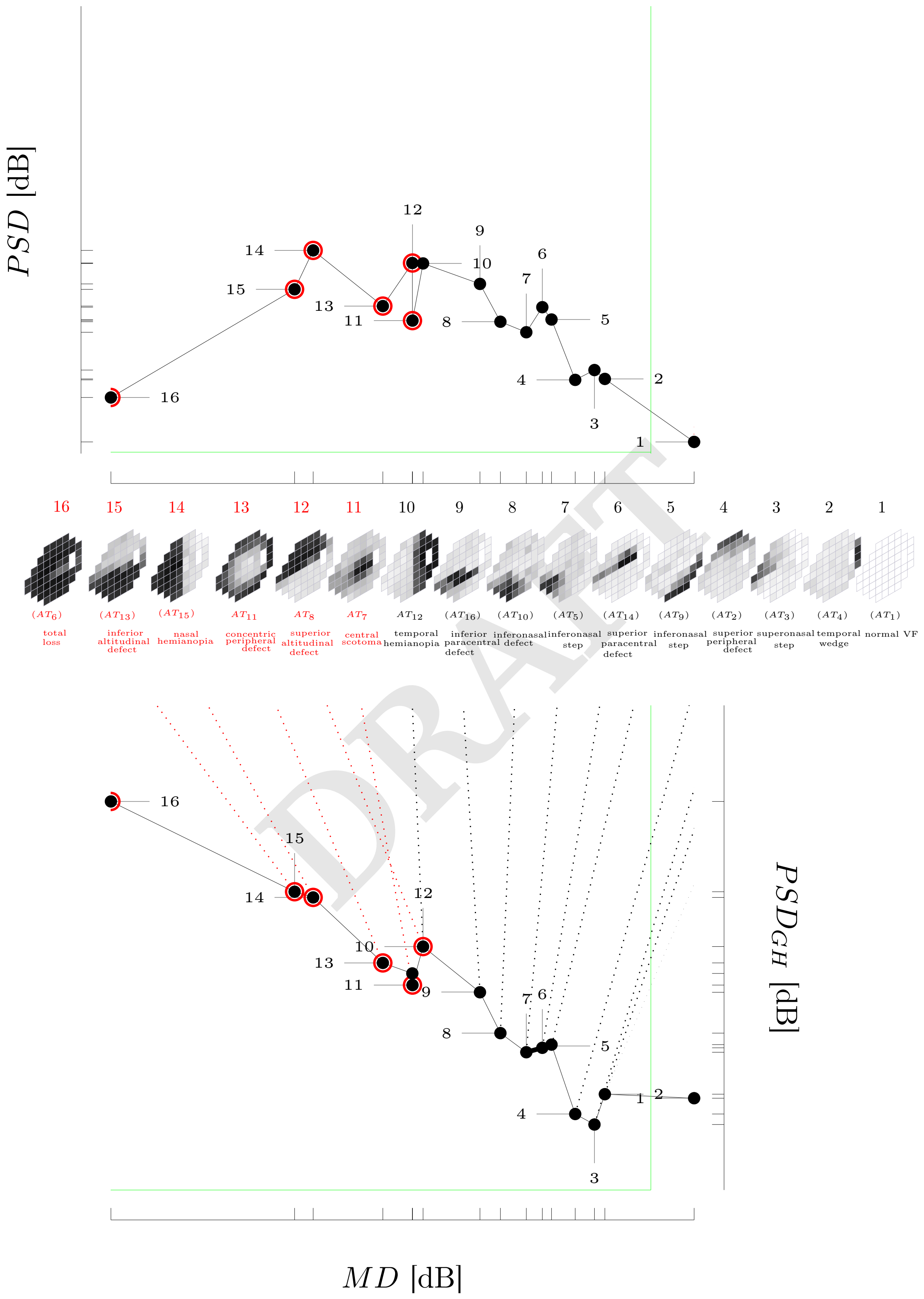


DRAFT

**Fig. 2.** The summary metrics MD and PSD from the 16 archtypes are plotted in two graphs with the pattern standard deviation calculated from the mean deviation obtained globally (upper graph) and locally (lower graph)(2). The first coordinate (MD) is identical in both graphs with the 2nd coordinate showing the effect of calculating the total average TD from the all the 52 test locations (upper graph) or local regions (lower graph) as seen below (next page)

.

DRAFT

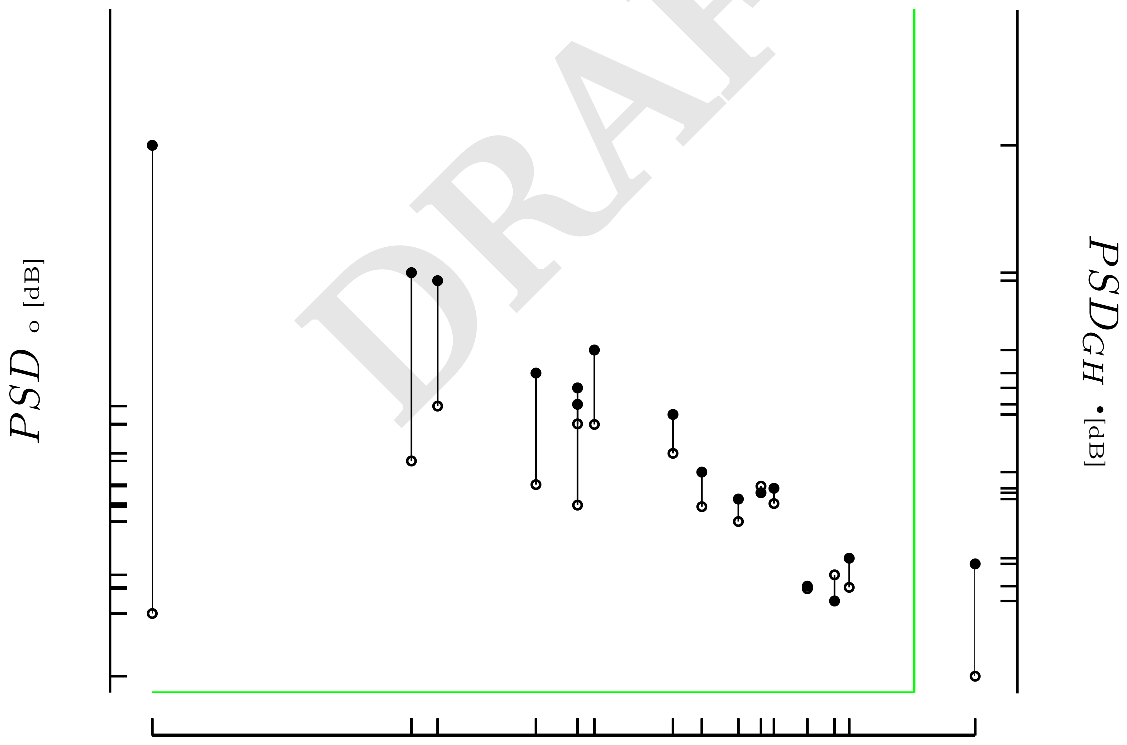


1

**Fig. 3.** The vertical line plot showing the difference between PSD$_{GH}$ and the PSD at the respective MD for the VF archtypes (1).

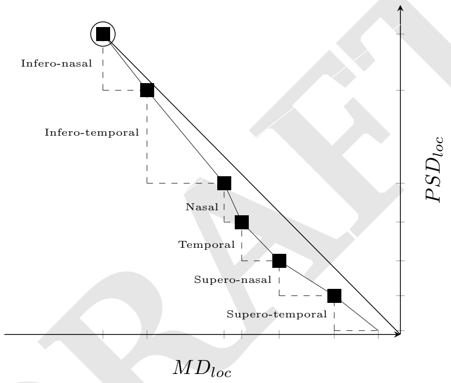


Figure 1: The cumulative proportions of a local mean and local pattern standard deviations [dB] $MD_{loc}$, $PSD_{loc}$ by sector (GH) for a individual VF.

DRAFT

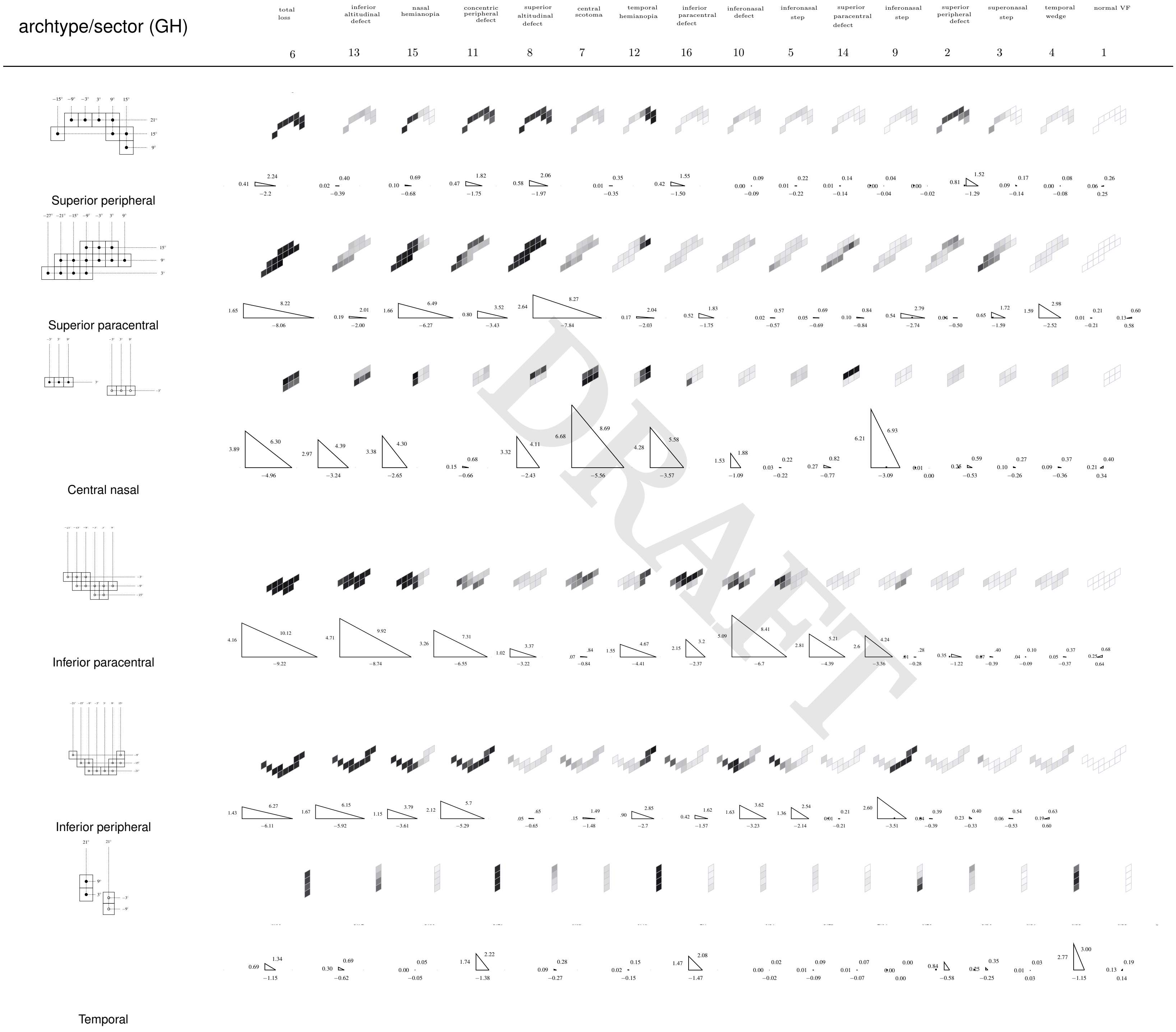


DRAFT

**Fig. 4.** The components MD$_{loc}$ and PSD$_{loc}$ by GH sectors for the VF archtypes (1)(3)

1. T. Elze, L. R. Pasquale, L. Q. Shen, T. C. Chen, J. L. Wiggs, and P. J. Bex. Patterns of functional vision loss in glaucoma determined with archetypal analysis. *J R Soc Interface*, 12(103), Feb 2015.
2. D. S. W. Ting, L. Peng, A. V. Varadarajan, P. A. Keane, P. M. Burlina, M. F. Chiang, L. Schmetterer, L. R. Pasquale, N. M. Bressler, D. R. Webster, M. Abramoff, and T. Y. Wong. Deep learning in ophthalmology: The technical and clinical considerations. *Prog Retin Eye Res*, Apr 2019.
3. D. F. Garway-Heath, D. Poinoosawmy, F. W. Fitzke, and R. A. Hitchings. Mapping the visual field to the optic disc in normal tension glaucoma eyes. *Ophthalmology*, 107(10):1809–1815, Oct 2000.

DRAFT
